# New onset COVID-19–related diabetes: an indicator of mortality

**DOI:** 10.1101/2020.04.08.20058040

**Authors:** Jin-Kui Yang, Jian-Min Jin, Shi Liu, Peng Bai, Wei He, Fei Wu, Xiao-Fang Liu, Zhong-Lin Chai, De-Min Han

## Abstract

**Background:** Concomitance with diabetes is associated with high mortality in critical conditions. Patients with previous diabetes are more vulnerable to COVID-19. However, new-onset COVID-19–related diabetes (CRD) and its relevance have scarcely been reported. This study investigates new-onset CRD and its correlation with poor outcomes or death in patients with COVID-19.

**Methods:** We performed a single center, retrospective case series study in 120 patients with laboratory confirmed COVID-19 at a university hospital. Fasting blood glucose (FBG) ≥7.0 mmol/L for two times during hospitalization and without a history of diabetes were defined as CRD. The *Critical* status was defined as admitted to intensive care unit (ICU) or death.

**Results:** After excluding patients with a history of diabetes, chronic heart, kidney, and liver disease, 69 patients with COVID-19 were included in the final analysis. Of the 69 patients, 23 were *Moderate*, 20 were *Severe*, and 26 were *Critical* (including 16 deceased patients). The prevalence of CRD in *Critical* and *Moderate+Severe* patients was 53.85% and 13.95%, respectively. Kaplan-Meier survival analysis revealed a significantly higher mortality rate in patients with CRD (P=0.0019). Multivariable analysis indicated that CRD was an independent predictor for death (HR = 3.75, 95% CI 1.26-11.15). Cluster analysis suggested that indicators for multi-organ injury were interdependent, and more proximities of FBG with indicators for multi-organ injury was present.

**Conclusion:** Our results suggest that new onset COVID-19–related diabetes is an indicator of multi-organ injury and predictor for poor outcomes and death in COVID- 19 patients. As it is easy to perform for clinical practices and even self-monitoring, glucose testing will be much helpful for predicting poor outcomes to facilitate appropriate intensive care in patients with COVID-19.

**Funding:** National Key Research and Development Program of China; The Beijing Science and Technology Project.

**Significance of this study:** *Evidence before this study:* Concomitance with diabetes is associated with high mortality in critical conditions. Patients with previous diabetes are more vulnerable to COVID-19. However, new-onset COVID-19–related diabetes (CRD) and its relevance have scarcely been reported. Recently, an international group of leading diabetes researchers participating in the CoviDIAB Project have established a global registry of patients with Covid-19–related diabetes (covidiab.e-dendrite.com).

*Added value of this study?:* New-onset diabetes in COVID-19 defined as CRD was investigated. Correlation between CRD and poor outcomes or death in patients with COVID-19 was found. About half of the *Critical* patients have new onset CRD. CRD is the representative of the clustered indicators of multi-organ injury and is the predictor for poor outcomes and death.

*How might these results change the focus of research or clinical practice?:* Our results suggest that new onset diabetes is an indicator of multi-organ injury and predictor for poor outcomes and death in COVID-19 patients. The study of CRD may also uncover novel mechanisms of disease.

## Introduction

In early December 2019, an outbreak of a novel coronavirus (SARS-CoV-2) pneumonia disease (COVID-19) occurred in Wuhan, China.[1] Most patients with COVID-19 were *Mild. Moderate* patients often experienced dyspnea after one week. Some *Severe* patients progressed rapidly to *Critical* condition including multi-organ failure and even death.[2-5]

Recently, new-onset diabetes in COVID-19 is noticed. An international group of leading diabetes researchers participating in the CoviDIAB Project have established a global registry of patients with Covid-19–related diabetes (covidiab.e-dendrite.com). [6] COVID-19 is caused by severe acute respiratory syndrome coronavirus 2 (SARS- CoV-2) infection. It is reminiscent of the SARS-CoV outbreak in early 2003, because both coronavirus attack cells via the same receptor, angiotensin-converting enzyme 2 (ACE2). [1]

We have previously reported that acute diabetes was commonly present in SARS patients without prior history of diabetes and without using glucocorticoids, and was an independent predictor for mortality in SARS patients.[7] Binding of SARS coronavirus to the angiotensin-converting enzyme 2 (ACE2) receptor may damage islets and causes acute diabetes.[8] *ACE2* gene knockout leads to diabetes in mice.[9]

ACE2 are expressed in key metabolic organs and tissues, including pancreatic islet cells,[10] the small intestine, adipose tissue, and the kidneys.[11] Thus, it is plausible that COVID-19 may have pleiotropic alterations of glucose metabolism or new type of disease.[6] In this observational cohort study, we aim to explore the correlation between COVID-19–related diabetes (CRD) and poor outcomes or death in patients with COVID-19.

## Methods

### Participants

This retrospective cohort study included consecutive COVID-19 adult inpatients (≥18 years old) admitted to Wuhan Union Hospital (Wuhan, China) and treated by the supportive medical team of Beijing Tongren Hospital (Beijing, China) from January 29, 2020, to March 20, 2020. A uniformed protocol of clinical practices was used in this cohort.

Respiratory specimens were collected by the local center for disease control and prevention (CDC) and then shipped to designated authoritative laboratories to detect SARS-CoV-2. The presence of SARS-CoV-2 in respiratory specimens was detected by real-time RT-PCR methods. The RT-PCR assay was conducted as per the protocol established by the World Health Organization (WHO).

The clinical outcomes including admitted to intensive care unit (ICU) or in-hospital death were monitored up to March 24, 2020, the final date of follow-up.

#### Exclusion criteria

(1) Patients with a history of diabetes were excluded. (2) To avoid glucocorticoid-induced diabetes, patients who received glucocorticoid treatment were also excluded. (3) For the identification of indicators of early multi-organ injury for predicting poor outcomes, patients with chronic organ damage, including heart disease (myocardial infarction and heart failure), kidney disease (maintenance dialysis or renal transplantation), or liver disease (liver cirrhosis) were excluded

#### COVID-19–related diabetes

FBG≥7.0 mmol/L for two times during hospitalization and without a history of diabetes in COVID-19 patients were defined as CRD

#### Clinical classification of COVID-19

Diagnosis and clinical classification criteria and treatment plan (version 6.0) of COVID-19 was launched by the National Health Committee of China (http://www.nhc.gov.cn/). The clinical classification of severity is as follows: (1) *Mild*, having only mild symptoms, imaging shows no pneumonia. (2) *Moderate*, with fever, respiratory tract symptoms, and imaging shows pneumonia. (3) *Severe*, meet any of the following signs: a) respiratory distress, respiratory rate ≥ 30 beats / min; b) in the resting state, finger oxygen saturation ≤ 93%) arterial blood oxygen partial pressure (PaO2/oxygen concentration (FiO2) ≤ 300mmHg (1mmHg = 0.133kPa). (4) *Critical*, one of the following conditions: a) respiratory failure occurs and requires mechanical ventilation; b) Shock occurs; c) ICU admission is required for combined organ failure

The Research Ethics Commission of Beijing Tongren Hospital, Capital Medical University (TRECKY2020-013) approved the study and the Ethics Commission waived the requirement for informed consent.

### Data collection

Demographic, clinical, laboratory and outcome data were extracted from the electronic hospital information system using a standardized form. All medical data were checked by two medical doctors (JMJ and PB) and the leader author (JKY) adjudicated any different interpretation between the two medical doctors.

### Statistical analysis

Data were expressed as median (interquartile range (IQR)) or percentage, as appropriate. Comparison of continuous data among different severity groups were determined using a Kruskal-Wallis test. Chi-square tests were used for categorical variables. Proximity Matrix of six measures as different organ damage indicators were calculated by Hierarchical cluster analysis. To find the risk factors predicting in-hospital death, univariate and multivariate Cox proportional hazard model were used to calculated hazard ratio (HR). Kaplan–Meier survival curves and the log-rank test were used for testing the survival between normal and abnormal indicators. SPSS for Windows 17.0 and Graphpad prism 7.0 software were used for statistical analysis, with statistical significance set at 2-sided P<0.05.

## Results

### Demographic and clinical characteristics

A total of 120 consecutive COVID-19 adult inpatients (≥18 years old) were admitted to Wuhan Union Hospital (Wuhan, China) and treated by the supportive medical team of Beijing Tongren Hospital (Beijing, China) from January 29, 2020 to March 20, 2020. Because Wuhan Union Hospital was assigned responsibility for the treatments of severe COVID-19 patients by the Wuhan government, most patients were *Severe* or *Critical*, some were *Moderate*, but none were *Mild*.

After excluding patients received glucocorticoid treatment or with a history of diabetes, myocardial infarction, heart failure, dialysis, renal transplant, cirrhosis, and patients missing basic medical information, 69 patients were included in the final analysis. Of the 69 patients, 23 were *Moderate*, 20 were *Severe*, and 26 were *Critical* (including 16 deceased patients). The median age of the 69 patients was 61 years (IQR, 52 to 67). Older patients tend to be more serious (p=0.024). The ratio of male to female patients were relatively similar, with 49.3% males. However, the proportion of males in deceased patients were 81.2%, suggestive that males may have higher mortality from COVID-19 (P=0.003). Fever (89.9), cough (65.2%) and dyspnea (43.5%) were the most common symptoms, while sputum (17.4%) and diarrhea (17.4%) were less common.

An increase in white blood cells and neutrophils, and decrease in lymphocytes and platelets occurred in *Critical* patients at the time of admission. A decrease of albumin, an increase of globulin, and an increase of C-reactive protein was also present in *Critical* patients (Table 1).

**Table 1:**
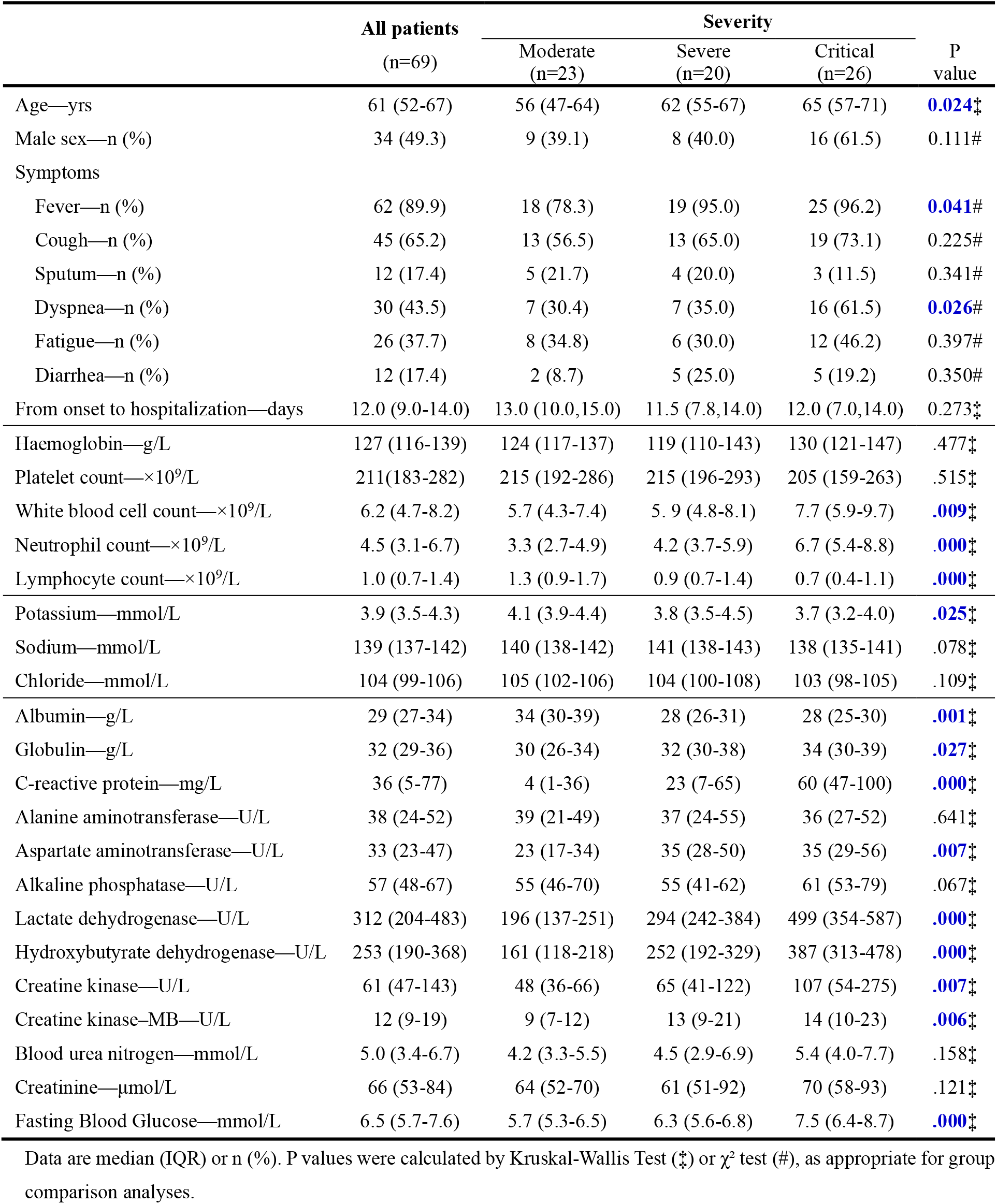
Demography, clinical and laboratory parameters of patients with COVID-19. Data are median (IQR) or n (%). P values were calculated by Kruskal-Wallis Test (‡) or χ^2^ test (#), as appropriate for group comparison analyses.

|  | All patients<br>(n=69) | Severity |  |  | P<br>value |
| --- | --- | --- | --- | --- | --- |
|  |  | Moderate<br>(n=23) | Severe<br>(n=20) | Critical<br>(n=26) |  |
| Age—yrs | 61 (52-67) | 56 (47-64) | 62 (55-67) | 65 (57-71) | **.024**‡ |
| Male sex—n (%) | 34 (49.3) | 9 (39.1) | 8 (40.0) | 16 (61.5) | 0.111# |
| Symptoms |  |  |  |  |  |
| Fever—n (%) | 62 (89.9) | 18 (78.3) | 19 (95.0) | 25 (96.2) | **.041**# |
| Cough—n (%) | 45 (65.2) | 13 (56.5) | 13 (65.0) | 19 (73.1) | 0.225# |
| Sputum—n (%) | 12 (17.4) | 5 (21.7) | 4 (20.0) | 3 (11.5) | 0.341# |
| Dyspnea—n (%) | 30 (43.5) | 7 (30.4) | 7 (35.0) | 16 (61.5) | **.026**# |
| Fatigue—n (%) | 26 (37.7) | 8 (34.8) | 6 (30.0) | 12 (46.2) | 0.397# |
| Diarrhea—n (%) | 12 (17.4) | 2 (8.7) | 5 (25.0) | 5 (19.2) | 0.350# |
| From onset to hospitalization—days | 12.0 (9.0-14.0) | 13.0 (10.0,15.0) | 11.5 (7.8,14.0) | 12.0 (7.0,14.0) | 0.273‡ |
| Haemoglobin—g/L | 127 (116-139) | 124 (117-137) | 119 (110-143) | 130 (121-147) | .477‡ |
| Platelet count—$\times 10^9/L$ | 211(183-282) | 215 (192-286) | 215 (196-293) | 205 (159-263) | .515‡ |
| White blood cell count—$\times 10^9/L$ | 6.2 (4.7-8.2) | 5.7 (4.3-7.4) | 5.9 (4.8-8.1) | 7.7 (5.9-9.7) | **.009**‡ |
| Neutrophil count—$\times 10^9/L$ | 4.5 (3.1-6.7) | 3.3 (2.7-4.9) | 4.2 (3.7-5.9) | 6.7 (5.4-8.8) | **.000**‡ |
| Lymphocyte count—$\times 10^9/L$ | 1.0 (0.7-1.4) | 1.3 (0.9-1.7) | 0.9 (0.7-1.4) | 0.7 (0.4-1.1) | **.000**‡ |
| Potassium—mmol/L | 3.9 (3.5-4.3) | 4.1 (3.9-4.4) | 3.8 (3.5-4.5) | 3.7 (3.2-4.0) | **.025**‡ |
| Sodium—mmol/L | 139 (137-142) | 140 (138-142) | 141 (138-143) | 138 (135-141) | .078‡ |
| Chloride—mmol/L | 104 (99-106) | 105 (102-106) | 104 (100-108) | 103 (98-105) | .109‡ |
| Albumin—g/L | 29 (27-34) | 34 (30-39) | 28 (26-31) | 28 (25-30) | **.001**‡ |
| Globulin—g/L | 32 (29-36) | 30 (26-34) | 32 (30-38) | 34 (30-39) | **.027**‡ |
| C-reactive protein—mg/L | 36 (5-77) | 4 (1-36) | 23 (7-65) | 60 (47-100) | **.000**‡ |
| Alanine aminotransferase—U/L | 38 (24-52) | 39 (21-49) | 37 (24-55) | 36 (27-52) | .641‡ |
| Aspartate aminotransferase—U/L | 33 (23-47) | 23 (17-34) | 35 (28-50) | 35 (29-56) | **.007**‡ |
| Alkaline phosphatase—U/L | 57 (48-67) | 55 (46-70) | 55 (41-62) | 61 (53-79) | .067‡ |
| Lactate dehydrogenase—U/L | 312 (204-483) | 196 (137-251) | 294 (242-384) | 499 (354-587) | **.000**‡ |
| Hydroxybutyrate dehydrogenase—U/L | 253 (190-368) | 161 (118-218) | 252 (192-329) | 387 (313-478) | **.000**‡ |
| Creatine kinase—U/L | 61 (47-143) | 48 (36-66) | 65 (41-122) | 107 (54-275) | **.007**‡ |
| Creatine kinase—MB—U/L | 12 (9-19) | 9 (7-12) | 13 (9-21) | 14 (10-23) | **.006**‡ |
| Blood urea nitrogen—mmol/L | 5.0 (3.4-6.7) | 4.2 (3.3-5.5) | 4.5 (2.9-6.9) | 5.4 (4.0-7.7) | .158‡ |
| Creatinine—$\mu\text{mol/L}$ | 66 (53-84) | 64 (52-70) | 61 (51-92) | 70 (58-93) | .121‡ |
| Fasting Blood Glucose—mmol/L | 6.5 (5.7-7.6) | 5.7 (5.3-6.5) | 6.3 (5.6-6.8) | 7.5 (6.4-8.7) | **.000**‡ |

Data are median (IQR) or n (%). P values were calculated by Kruskal-Wallis Test (‡) or $\chi^2$ test (#), as appropriate for group comparison analyses.

### COVID-19–related diabetes and mortality

The prevalence of CRD in *Critical* and *Moderate+Severe* patients was 53.85% and 13.95%, respectively (Figure 1 A and B). Kaplan-Meier survival analysis revealed a significantly higher mortality rate in patients with CRD (P=0.0019) (Figure 1C).

**Figure 1.**
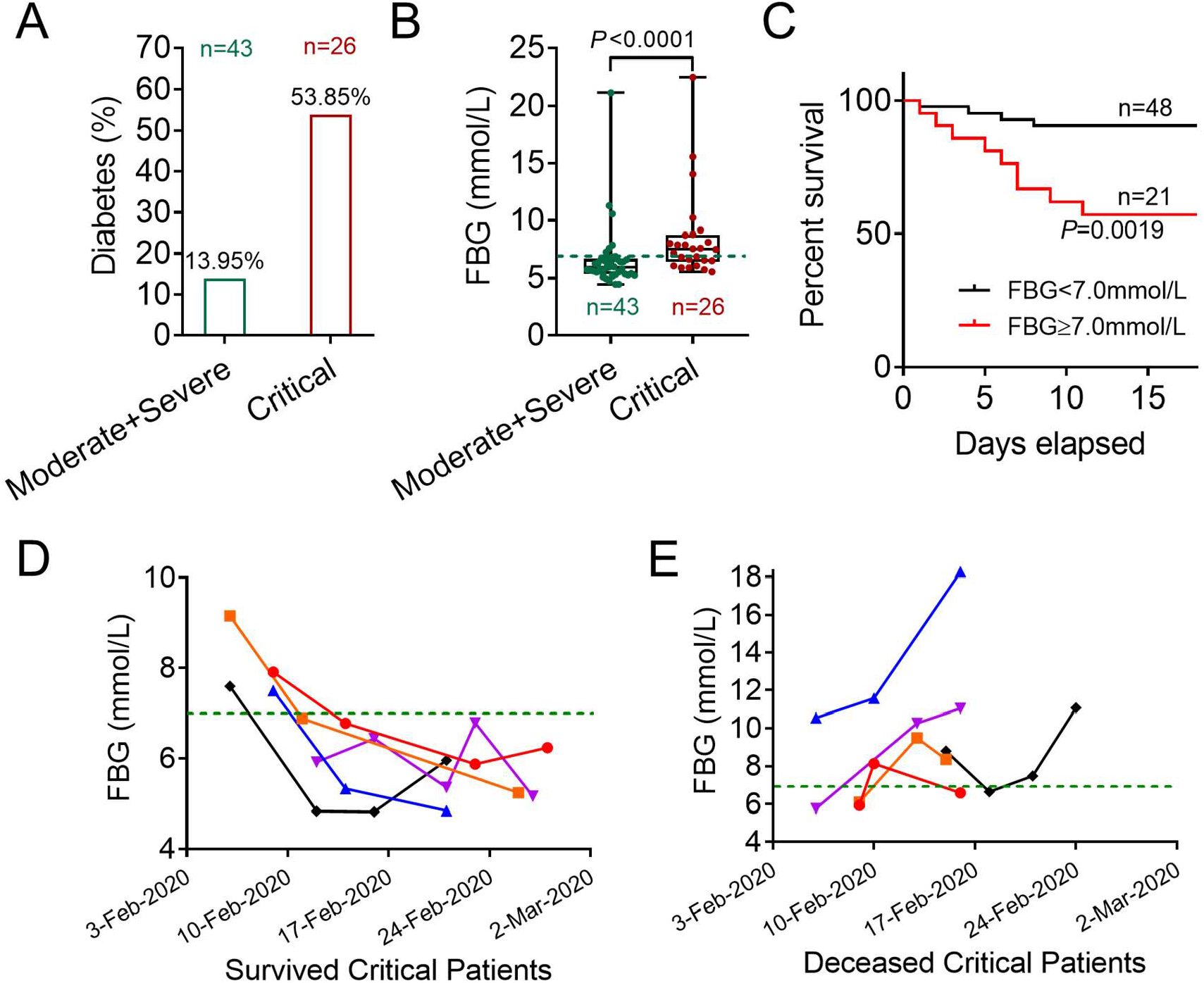
COVID-19–related diabetes and mortality. **(A)** Prevalence of COVID-19–related diabetes in *Critical* and *Moderate+Severe* groups. **(B)**Fasting blood glucose levels in *Critical* and *Moderate+Severe* groups. **(C)**Kaplan–Meier survival curves for in-hospital death rate of patients with COVID- 19. **(D-E)** Longitudinal observation of Fasting blood glucose during admission and subsequent treatment.

### Longitudinal changes of FBG and mortality

FBG level was much higher above the criteria of diagnosis of diabetes in *Critical* group, while it did not increase remarkably in *Moderate+Severe* group (Figure 1B). Considering that the deaths happened only in *Critical* patients in this cohort, a longitudinal case-control investigation was conducted in *Critical* patients. In 26 *Critical* patients who were measured once every three days and at least three times were analyzed, while those who did not have >3 measurements were not included for analysis. Although FBG levels were much higher above the upper limit of normal value at the time of admission, they dramatically decreased to the normal range during subsequent treatment in survivors. However, of the patients who died, increases of FBG levels continued (Figure 1D-E).

### FBG as a predictor for multi-organ injury and mortality

Lactate dehydrogenase (LDH)/hydroxybutyrate dehydrogenase (HBDH), creatinine (Cr), and alanine aminotransferase (ALT) were used as the relative function of multiple organs (i.e., the heart, kidney, and liver function, respectively). Cluster analysis suggested that some definite indicators for multi-organ injury were independent and that multi-organ injury caused by the virus infection might present. Most interestingly, significant proximities or interdependent of FBG with other Variables including LDH (*r*=0.433, *P*<0.01), HDDH (*r*=0.43, *P*<0.01), and Cr (*r*=0.498, *P*<0.01) were present. (Table 2).

**Table 2:** Proximity matrix of six multi-organ injury indicators by Hierarchical cluster analysis. FBG: Fasting Blood Glucose; LDH: Lactate dehydrogenase; Cr: Creatinine; CK: Creatine kinase; HDDH: Hydroxybutyrate dehydrogenase; ALT: Alanine aminotransferase.

|  | **FBG** | **LDH** | **Cr** | **CK** | **HDDH** | **ALT** |
| --- | --- | --- | --- | --- | --- | --- |
| **FBG** | 1 |  |  |  |  |  |
| **LDH** | **\*\*0.433** | 1 |  |  |  |  |
| **Cr** | **\*\*0.498** | 0.265 | 1 |  |  |  |
| **CK** | 0.025 | *0.328 | 0.113 | 1 |  |  |
| **HDDH** | **\*\*0.428** | **\*\*0.881** | 0.128 | **\*\*0.385** | 1 |  |
| **ALT** | 0.099 | **\*\*0.441** | 0.258 | 0.017 | 0.246 | 1 |

\*\* p<0.01; \* P<0.05. Hierarchical cluster analysis, Pearson correlation test (2-tailed).

Using the univariable Cox proportional hazards model, male sex (HR = 5.02, 95% CI 1.43-17.62), elevated FBG (HR = 5.09, 95% CI 1.76-14.70), LDH (HR = 9.73, 95% CI 1.28-73.7) and HBDH (HR = 3.81, 95% CI 1.23-11.85) were predictors for death (Figure 2A). In the multivariable model, elevated FBG remained the independent predictor for death (HR = 3.75, 95% CI 1.26-11.15) (Figure 2B).

**Figure 2.**
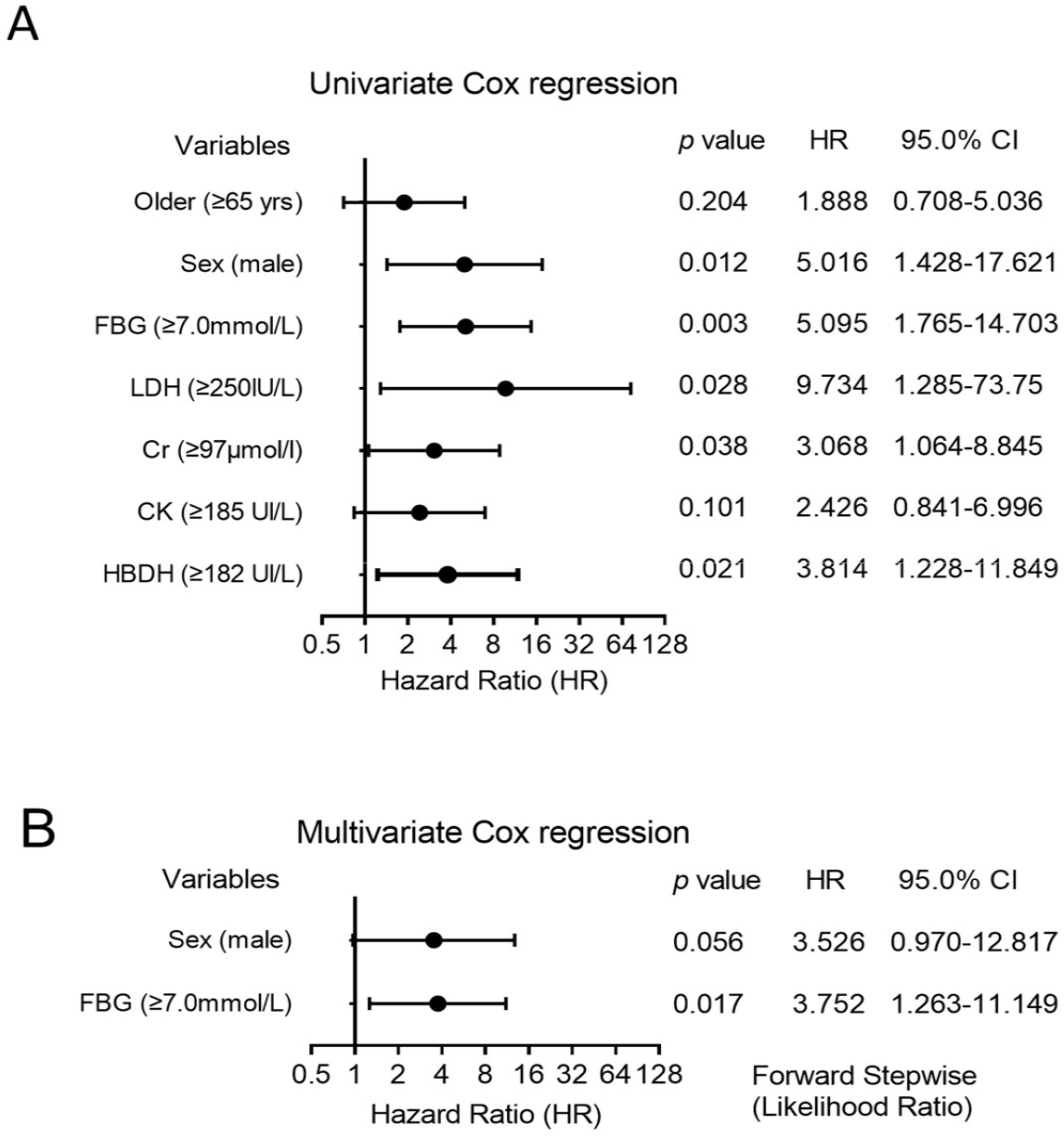
Effects of multi-organ injury indicators on the risk of in-hospital death in patients with COVID-19. **(A)** Univariable Cox regression indicated effects of multi-organ injury indicators on predicting in-hospital death. **(B)** Multivariable Cox regression indicated effects of independent multi-organ injury indicators on predicting in-hospital death.

## Discussion

This study has shown that new-onset diabetes is very common in COVID-19. The prevalence of CRD in *Critical* and *Moderate+Severe* patients was 53.85% and 13.95%, respectively. The results also suggest that CRD is an indicator of multi-organ injury and predictor for poor outcomes and death in COVID-19 patients.

Except pneumonia, the multi-organ nature of COVID-19 or SARS-CoV-2 infection has been demonstrated in a latest autopsy studies.[12] Moreover, an autopsy study on 3 cases of SARS indicate that some lesions observed in heart, kidney, pancreas may have existed before the hospitalization.[13] We found that ACE2, the SARS-CoV-2 receptor, is highly expressed in the pancreatic islet.[10]

Subacute islet injury caused by virus infection has been widely reported in type 1 diabetes, which is an autoimmune disease characterized by a long-term loss of pancreatic islet β-cells. Acute pancreatic islet injury induced diabetes caused by virus infection has scarcely been reported. However, serological evidence of infection and isolation of viruses including Coxsackie B and Mumps from the pancreas have been reported in a few cases of acute diabetes.[14] A case-control study indicated that high FBG is an independent predictor for severity of H1N1 pneumonia. In the study, FBG was remarkably increased in patients with H1N1 pneumonia than in patients with non- H1N1 pneumonia (8.3 vs. 6.2 mmol/L). Moreover, compensative rise in insulin, in corresponding with high FBG was not found; instated, islet β-cells function indicated by HOMA-β index is lower (103 vs. 134).[15] We have previously reported that hyperglycemia is an independent predictor for death in SARS patients.[7] Binding of SARS coronavirus to the ACE2 receptor may lead to impairment of the function of ACE2 and the functional loss of ACE2, as observed in the ACE2 KO mice, is known to cause diabetes.[9] Furthermore, coronavirus can use the ACE2 on the islet cells as its receptor to enter and replicate within the islet cells, leading to damage of islet cells, such as insulin producing β cells, ultimately leading to insulin deficiency, a direct cause of acute diabetes, as observed in SARS patients.[8]

It is appreciated that FBG elevation can be a result of “Stress hyperglycemia” during acute illness in patients with COVID-19. However, severe hyperglycemia can also be caused by either insulin deficiency or insulin resistance. Insulin resistance caused by metabolic, hormonal, and cytokine changes as a result of illness is usually associated with a corresponding rise in insulin output, which is a compensation mechanism to maintain normal glycemia. Only when the insulin resistance was too severe and the compensation failed, the level of blood glucose would increase significantly [16, 17]. Therefore, relative dysfunction of pancreatic islet insulin secretion caused by the virus infection could be a main reason for the severe hyperglycemia observed in patients with COVID-19.

Our results suggest that increased level of blood glucose is a representative of the clustered multi-organ injury indicators and an earlier predictor for poor outcomes, including death in these patients. Acute heart injury such as acute myocarditis is common in patients with infection of various viruses, such as adenovirus, herpesvirus and enterovirus [18]. Since myocarditis associated with coronavirus infection was firstly reported in 1980 [19], a growing body of evidence shows that coronavirus is also a pathogen for heart injury that should not be ignored. [8, 20] The latest case report has described cardiac injury in a patient with COVID-19.[21] Two myocardial enzymes, LDH and HBDH are known to be released into the circulation as a result of the myocardial injury. In this study, we found that high levels of both LDH and HBDH in patients on admission were predictor for severity and in-hospital death. Acute kidney injury is also common in patients with virus infection, such as Parvovirus B19, Hanta, Ebola, and Dengue virus infection.[22] Coronavirus associated kidney injury was reported in patients with MDRS.[23] In the previous report, we found that SARS-CoV may cause kidney injury and elevated Cr is an independent predictor for in-hospital death in SARS patients.[8] In this study, we found high Cr were predictor for in-hospital death in COVID-19 patients.

There are several limitations in our study. First, we only assessed FBG and some most accessible biochemical parameters. It would have produced better results if some more specific indicators such as serum insulin levels, echocardiography, glomerular filtration rate were measured in this study. Second, interpretation might be limited by the sample size of the study. Last but not least, missing of HbA1c level is a limitation. Because, low HbA1c would indicate no history of recent diabetes and support the confirmation of new onset of diabetes.

In summary, except pneumonia, multi-organ injury including the heart, kidney, and possibly pancreatic islet injuries can cause CRD at an early stage and thereby increase the risk of mortality in patients with COVID-19, if hyperglycemia cannot be improved as shown in Fig 1. These multi-organ injury indicators were associated with higher odds of death and were interdependent, indicating that the multi-organ damage happened at the same time. Among them, COVID-19–related diabetes is a representative of the clustered multi-organ injury for predicting mortality of COVID-19. As it is easy to perform for clinical practices and self-monitoring, blood glucose testing will be much helpful for predicting critical condition to facilitate appropriate intensive care.

## Data Availability

The data are available from the correspondence author

## Contributions

JMJ, PB, FG, WH, SL, FW, DMH SL and JKY collected the epidemiological and clinical data and processed statistical data. JKY drafted the manuscript. JKY, JMJ, SL and CZL revised the final manuscript. JKY is responsible for summarizing all epidemiological and clinical data.

## Acknowledgments

This study was funded by the National Key R&D Program of China (2017YFC0909600). The funder has no role in study design, data collection, data analysis, manuscript preparation and/or publication decisions. We thank all patients involved in the study.

## References

1. Zhu N, Zhang D, Wang W, Li X, Yang B, Song J, et al. A Novel Coronavirus from Patients with Pneumonia in China, 2019. N Engl J Med 2020.

2. Huang C, Wang Y, Li X, Ren L, Zhao J, Hu Y, et al. Clinical features of patients infected with 2019 novel coronavirus in Wuhan, China. Lancet 2020; 395:497–506.

3. Chen N, Zhou M, Dong X, Qu J, Gong F, Han Y, et al. Epidemiological and clinical characteristics of 99 cases of 2019 novel coronavirus pneumonia in Wuhan, China: a descriptive study. Lancet 2020; 395:507–513.

4. Wang D, Hu B, Hu C, Zhu F, Liu X, Zhang J, et al. Clinical Characteristics of 138 Hospitalized Patients With 2019 Novel Coronavirus-Infected Pneumonia in Wuhan, China. JAMA 2020.

5. Xu XW, Wu XX, Jiang XG, Xu KJ, Ying LJ, Ma CL, et al. Clinical findings in a group of patients infected with the 2019 novel coronavirus (SARS-Cov-2) outside of Wuhan, China: retrospective case series. BMJ 2020; 368:m606.

6. Rubino F, Amiel SA, Zimmet P, Alberti G, Bornstein S, Eckel RH, et al. New-Onset Diabetes in Covid-19. N Engl J Med 2020.

7. Yang JK, Feng Y, Yuan MY, Yuan SY, Fu HJ, Wu BY, et al. Plasma glucose levels and diabetes are independent predictors for mortality and morbidity in patients with SARS. Diabet Med 2006; 23:623–628.

8. Yang JK, Lin SS, Ji XJ, Guo LM. Binding of SARS coronavirus to its receptor damages islets and causes acute diabetes. Acta Diabetol 2010; 47:193–199.

9. Niu MJ, Yang JK, Lin SS, Ji XJ, Guo LM. Loss of angiotensin-converting enzyme 2 leads to impaired glucose homeostasis in mice. Endocrine 2008; 34:56–61.

10. Fang HJ, Yang JK. Tissue-specific pattern of angiotensin-converting enzyme 2 expression in rat pancreas. J Int Med Res 2010; 38:558–569.

11. Hamming I, Timens W, Bulthuis ML, Lely AT, Navis G, van Goor H. Tissue distribution of ACE2 protein, the functional receptor for SARS coronavirus. A first step in understanding SARS pathogenesis. J Pathol 2004; 203:631–637.

12. Yao XH, Li TY, He ZC, Ping YF, Liu HW, Yu SC, et al. [A pathological report of three COVID-19 cases by minimally invasive autopsies]. Zhonghua Bing Li Xue Za Zhi 2020; 49:E009.

13. Lang Z, Zhang L, Zhang S, Meng X, Li J, Song C, et al. Pathological study on severe acute respiratory syndrome. Chin Med J (Engl) 2003; 116:976–980.

14. Jaeckel E, Manns M, Von Herrath M. Viruses and diabetes. Ann N Y Acad Sci 2002; 958:7–25.

15. Wang W, Chen H, Li Q, Qiu B, Wang J, Sun X, et al. Fasting plasma glucose is an independent predictor for severity of H1N1 pneumonia. BMC Infect Dis 2011; 11:104.

16. Gupta D, Krueger CB, Lastra G. Over-nutrition, obesity and insulin resistance in the development of beta-cell dysfunction. Curr Diabetes Rev 2012; 8:76–83.

17. Dungan KM, Braithwaite SS, Preiser JC. Stress hyperglycaemia. Lancet 2009; 373:1798–1807.

18. Pollack A, Kontorovich AR, Fuster V, Dec GW. Viral myocarditis--diagnosis, treatment options, and current controversies. Nat Rev Cardiol 2015; 12:670–680.

19. Riski H, Hovi T, Frick MH. Carditis associated with coronavirus infection. Lancet 1980; 2:100–101.

20. Alhogbani T. Acute myocarditis associated with novel Middle east respiratory syndrome coronavirus. Ann Saudi Med 2016; 36:78–80.

21. Inciardi RM, Lupi L, Zaccone G, Italia L, Raffo M, Tomasoni D, et al. Cardiac Involvement in a Patient With Coronavirus Disease 2019 (COVID-19). JAMA Cardiol 2020.

22. Prasad N, Novak JE, Patel MR. Kidney Diseases Associated With Parvovirus B19, Hanta, Ebola, and Dengue Virus Infection: A Brief Review. Adv Chronic Kidney Dis 2019; 26:207–219.

23. Mackay IM, Arden KE. MERS coronavirus: diagnostics, epidemiology and transmission. Virol J 2015; 12:222.

